# Variations in housing affordability and health relationships by measurement and sub-population assessment

**DOI:** 10.1101/2021.02.03.21251094

**Authors:** Will Raderman, Nina T. Rogers, Emily T. Murray

## Abstract

Past studies have shown that the more unaffordable housing is to people, the worse their health, particularly mental health. However, the commonly used housing affordability indicator, the 30% measure, has limitations. There is evidence that other indicators, including the ‘30/40’ measure, might be more precise in characterizing housing unaffordability by taking into account absolute values of household incomes. In this paper, we use cross-sectional data from the UK Household Longitudinal Study, to evaluate relationships between two affordability measures (30%, 30/40) with 3 health measures: general, physical and mental health. We use logistic regression and effect modification to test whether relationships varied by age, ethnicity, housing tenure, urbanicity and sex. Out of 35,114 participants with complete data, housing was classified as unaffordable for 24.2% using the 30% measure and 10.2% for the 30/40 measure. In age-adjusted analyses, higher unaffordability was associated with worse health for all three health measures, with associations stronger for the 30/40 vs the 30% unaffordability measure. In models adjusted for age, sex and urbanicity, both ethnicity and tenure independently modified associations; with modification showing small differences by unaffordability and health measure. Further studies are needed to disentangle complex relationships between household income, housing costs, ethnicity and tenure.

## Introduction

Housing is a basic human need. In the same way that quality food, clean air, and exercise are essential to good health, housing heavily dictates the wellbeing of people (Matte & Jacobs, 2000). Housing is an unavoidable cost for renters and those with a mortgage. Most individuals and families dedicate a significant portion of their earnings to pay for their living space. For example, houses in the UK cost nearly eight times more than average earnings in the UK (Chu, 2018) and Londoners spend 42% of pre-tax income on rent (Chapman, 2019). Costly housing is a major problem because individuals and families often face poor housing conditions, are less able to pay for essential goods and services (Stone, 2004), and as a result suffer poor health consequences, including cardiac and lung diseases (Ormandy & Ezratty, 2016) and reduced mental wellbeing (Akbar & Hossain, 2017; Jones-Rounds, Evans, & Braubach, 2014).

Previous studies have found associations between housing affordability and both mental (Cairney & Boyle, 2004; Mason et al, 2013; Clair & Hughes, 2019) and physical health (Ormandy & Ezratty, 2016; Fisher et al, 2009). A longitudinal study of The Household, Income and Labour Dynamics in Australia survey, which analyzed changes in the 36-item Short Form (SF-36) Mental Health Component Summary scores of adults, indicated housing affordability had more impact on mental health than general financial difficulties (Mason et al, 2013). Cairney and Boyle evaluated mental health through a questionnaire, finding that homeowners had less psychological distress than renters when adjusting for income level (Cairney & Boyle, 2004). Clair A et al. analyzed C-reactive protein (CRP), known for its connection to mental stress and acute infection, discovering an association between elevated CRP levels with housing unaffordability while controlling for demographic characteristics (Clair & Hughes, 2019).

Other studies have used indirect measures of housing unaffordability. Ormandy et al. analyzed the association with a composite socioeconomic status measure as a proxy for unaffordable housing (Ormandy & Ezratty, 2016). Excess winter mortality and thermal discomfort were related to harmful housing conditions and inadequate financial resources (Ormandy & Ezratty, 2016). In Fisher et al., an affordability index based on location and amenities was developed (Fisher et al, 2009). A correlation between the quality of town and affordability was discovered, where lower income households were dispersed to locations with poorer conditions (Fisher et al, 2009).

Different pathways are hypothesized to link housing unaffordability to poor mental health. Most directly, psychological distress is caused by elevated financial strain (Mason et al, 2013). Studies have shown that people who cannot afford to pay a higher percentage of their income on housing are more likely to live in poor quality housing (Baker et al, 2016). A review of 38 studies found that quality of housing is associated with poor mental health outcomes (i.e. clinical depression) (Baker et al, 2016). Housing with low accessibility to jobs results in higher travel costs and labor instability (Akbar & Hossain, 2017), leading to higher distress. Better quality neighborhoods and social networks attenuate the detrimental health effects of substandard housing. Neighborhoods with insecure environments reduce the opportunity for better mental states (Jones-Rounds et al, 2014). Families then rely more on their living space, leading to less social cohesion and higher emotional stress (Akbar & Hossain, 2017).

Mechanisms from housing unaffordability to physical health include the inability to afford adequate housing forcing many to cheaper, more isolated, and hazardous living spaces, leading to worse physical health (Fisher et al, 2009) and excess mortality (Ormandy & Ezratty, 2016). Unaffordable housing may also leave households less secure for other essential items like food (Stone, 2004), which results in worse physical health outcomes (Gupta et al, 2007).

Understanding the scale and specifics as to who is suffering health difficulties because of housing affordability issues is complicated (Stephen & Hoskara, 2019). The first challenge is how to accurately capture who has and has not experienced housing unaffordability. Traditionally, the proportion of income spent, specifically over or under 30%, on housing has been used to determine which households are experiencing unaffordable housing (Gabriel, 2005; Stone et al., 2011a; Yates, 2008). However, there are questions about whether the financial measure of 30% of income on housing is capturing the most vulnerable populations, since low income households spending less than 30% of income on housing may still be unable to meet all basic needs (Stone, 2004; Yates, 2008). Furthermore, there is minimal analysis comparing the demographics of a population according to different classifications of housing affordability.

In addition, past studies have identified sub-populations that experience greater risk of housing unaffordability. Younger individuals, particularly first-time buyers, are more prone to extreme cost burdens in OECD nations (Gan & Hill, 2009; OECD, 2011a). Renters, especially those of lower income, experience poorer mental health and higher levels of psychological distress than homeowners (Cairney & Boyle, 2004). Women and people of color experience significant rates of housing unaffordability. Female lone-parents and women living alone experience significant risk of being above the 30% cost to income ratio (Rea, 2008). White households have a lower shelter poverty rate than people of color (Stone, 2004). Black households are the least likely to own housing and most likely to be a social renter in the UK (Barton, 2017).

Yet complex relationships exist between sociodemographic indicators, housing affordability and health. Women, in particular have faced a number of health issues and disparities stemming from declining housing conditions (Pevalin et al, 2008). Younger age groups have also been found to be negatively impacted from the burden of high housing costs. Adolescents living in poor conditions suffer greater psychological distress, independent of income level (Baker et al, 2016). Children in overcrowded homes experience developmental delays and emotional problems, in addition to worse educational retention and mental adaptations (Gan & Hill, 2009). Therefore, it is unclear whether certain sub-population’s health are more or less vulnerable to the effects of housing unaffordability.

The aim of this study was therefore to assess whether housing affordability is associated with physical and mental health of adult participants of the Understanding Society study (Lynn, 2009), and whether these associations vary by affordability measures and demographic factors.

## Background

### Brief History of Housing Affordability in the UK Context

Many policy-makers and researchers believe that housing affordability issues in the UK started when Margaret Thatcher began the process of UK housing privatization over the past few decades with her ‘right to buy’ scheme (Norris & Byrne, 2018). It was thought that the wellbeing and financial status of citizens would be improved by increasing homeownership through privatization (Norris & Byrne, 2018). However, housing affordability declined in the UK as housing has become a commodity (McCord, 2013; Norris & Byrne, 2018).

Thatcher’s policy changes included selling government-controlled social housing and deregulation, where constraints on mortgage distribution were eliminated (McCord, 2013; Norris & Byrne, 2018). As demand for purchasing homes went up, so did the prices (Mulliner & Maliene, 2013). From the late 1970s onward, the government privatized housing and stopped constructing new social dwellings (Parvin et al, 2011). Prices rose drastically (Parvin et al, 2011), income levels did not rise accordingly (McCord, 2013). House price-to-income measures have steadily increased, going from below 3:1 in the early 1990s to a peak of 7.2:1 in late 2007 (McCord, 2013). Supply of housing did not match the rapid increase in demand, in part due to restrictive land use policies and the lack of new social housing developments, which heavily contributed to the large amount of market volatility and the United Kingdom’s housing crisis (Gan & Hill, 2009; OECD, 2011; Mulliner & Maliene, 2013).

First time buyers and lower income individuals were particularly hurt the most by these conditions (Czischke & van Bortel, 2018). At the same time, housing affordability was reduced for renters through rises in rental prices (McCord, 2013; Gan & Hill, 2009). After the 2008 housing crash, mortgage-lending criteria became stricter, with home purchasers required to put down high deposits and low loan-to-value ratios, difficult costs and barriers for those on lower incomes (McCord, 2013). The UK has also implemented austerity measures since the late 2000s, which have diminished disposable income levels (McCord, 2013). The reduction of government involvement in housing, both in terms of regulating and spending, has resulted in increased housing affordability issues for many (McCord, 2013; Mulliner & Maliene, 2013).

## Methods

### Data Source

Our study was comprised of participants of Understanding Society (US), a longitudinal study, tracking individuals from households resident in the United Kingdom (Lynn, 2009). The survey data, which includes an ethnic minority boost sample, is available through the UK Data Service (University of Essex, 2020). Information was collected primarily face-to-face, with a small number of interviews done by phone. This study was based on the baseline wave undertaken from 2009 to 2011.

### Housing affordability exposures

Two affordability measures were used in this analysis, spending equal to or more than: 30% of income on housing and 30% of income for people in the bottom 40% of the equivalised income distribution (Horsfield, 2015). Each exposure variable was calculated using household income and housing expenditures data, either rental or mortgage costs. Household income is an imputed measure from individual responses in each household. Income and housing expenditures were converted to weekly rates and adjusted for household size according to the modified OECD equivalence scale (OECD, 2011b). Each variable was coded into two categories of affordable and unaffordable.

### Physical and mental health outcomes

Three dimensions of health were evaluated using three indicators: the SF-12 physical composite score (PCS) for physical health, the SF-12 mental composite score (MCS) for mental health and self-rated health for an overall measure of general health. The SF-12 scores are derived from twelve questions in the US survey, taken from the larger SF-36 Health Survey (Gandek et al., 2004). The questions are scored on a 0 to 100 scale for mental and physical function, with 100 representing the best health ranking. General health was determined by a survey question asking participants whether their health was generally excellent, very good, good, fair, or poor.

### Covariates tested for effect modification

Based on the literature, sex, ethnicity, urbanicity, housing tenure, and age were selected as covariates for the analysis (Pevalin et al, 2008; Stone, 2004; Fisher et al, 2009; Cairney & Boyle, 2004; Baker et al, 2016). Participants had their sex coded as male or female. Age at the time of the survey was treated as a continuous covariate. Households were classified as either urban or rural in location based on a population of either more or less than 10,000 people, respectively. The covariate of ethnicity was collapsed from one category per specific nationality (n=18) to four groupings of similar ethnicities (Caucasian, Black, Asian, and Other categories). Those listed as Other ethnicity did not classify in the main three categories. The covariate of housing tenure was categorized as: full ownership, owned with mortgage, social rental (local authority, housing authority), private rental (rented private furnished, rented private unfurnished), and other (rented from employer, other). These re-categorizations were to maintain sufficient sample sizes in sub-group analyses.

## Statistical Methods

Analysis was carried out on a complete-case sample. Characteristics of the analytical sample were observed and numbers of participants in each category for affordability exposures, health outcomes, and covariates were noted. Mantel-Haenszel and Wilcoxon Rank Sum Tests were made between complete and incomplete samples’ distribution of all variables used.

The distributions of physical and mental health scores were skewed left, so SF-12 outcomes were converted to percent quartiles. Initially, age-adjusted logistic regression models were used to determine whether there were ordinal associations between the financial metrics (30% and 30/40 Rule) and physical, mental and general health. For regressions analyzed, the SF-12 quartiles were numbered in reverse order, so odds ratios should be interpreted as each 1-quartile increase in worse health associated with change in odds of unaffordability.

Second, relationships between housing affordability and health outcomes were tested for effect modification by sex, urbanicity, ethnicity and tenure. Initially, while controlling for age, logistic regressions were fitted between each affordability measure and health outcome with an interaction term added separately for each potential modifier. The final step of the analysis involved comprehensive logistic regressions between each housing affordability metric with each health outcome, with all significant interaction terms fitted in one model. If an interaction term p-value was <0.05, regressions were fitted separately for each modifier category. If effect modification was not indicated in singular models, covariates were included as potential confounders.

## Results

### Characteristics of the sample

Of the entire sample population totaling 77,309, there were 21,350 (27.6%) participants excluded for missing information related to housing payments. The remaining sample of 55,959 had no missing income values. An additional 20,644 people had incomplete covariate information. Lastly, 201 individuals were removed from the study sample for missing all health outcomes. This resulted in a final sample size of 35,114 (45.4 % of total) participants with complete financial, covariate, and health information.

The majority of the complete-case sample was female (56.1%), Caucasian (83.3%), urban (77.6%), and homeowners (82.2%), with an average age of 48.6 years old (Table 1). Most participants ranked themselves as having either Excellent (17.40%) or Very good (31.80%) general health. The SF-12 measures for physical and mental health were skewed to the left, with 82.05% and 85.56% of participants occurring in the upper half of the score range (highest levels of good health), respectively. Percent quartiles were used as a result. Housing was classified as unaffordable for 24.2% of the sample if the 30% income measure was used, compared to 10.2% for the 30/40 measure. Using either the 30% income or 30/40 measure, ethnic minorities (vs Caucasian), urban residents (vs rural), tenures other than outright homeowners, younger study members and those with worse mental health were more likely to be classed as living in unaffordable housing. For physical and general health, participants with better health were more likely to be considered as living in unaffordable housing according to the 30% income measure. In contrast, participants in the worst physical and general health categories faced the most unaffordability according to the 30/40 metric.

**Table 1:** Characteristics of covariates, affordability measures, and health metrics, Understanding Society 2009-11.

|  | 30% measure |  |  |  | 30/40 measure |  |  |
| --- | --- | --- | --- | --- | --- | --- | --- |
|  | TOTAL<br>(n=35,114) | Unaffordable<br>(n=8,480) | Affordable<br>(n=26,634) | p-values for<br>difference | Unaffordable<br>(n=3,572) | Affordable<br>(n=31,542) | p-values for<br>difference |
| Female, % | 56.1 | 24.48 | 75.52 | 0.263 | 10.60 | 89.40 | 0.015 |
| Ethnicity, % |  |  |  | <0.001 |  |  | <0.001 |
| Caucasian | 83.3 | 21.22 | 78.78 |  | 8.02 | 91.98 |  |
| Black | 4.8 | 40.93 | 59.07 |  | 20.94 | 79.06 |  |
| Asian | 10.4 | 38.80 | 61.20 |  | 22.18 | 77.82 |  |
| Other | 1.5 | 37.78 | 62.22 |  | 17.41 | 82.59 |  |
| Urban, % | 77.6 | 26.00 | 74.00 | <0.001 | 11.45 | 88.55 | <0.001 |
| Tenure, % |  |  |  | <0.001 |  |  | <0.001 |
| Owned outright | 38.0 | 0.00 | 100.00 |  | 0.00 | 100.00 |  |
| Mortgage | 44.2 | 40.65 | 59.35 |  | 11.52 | 88.48 |  |
| Social rental | 13.0 | 33.20 | 66.80 |  | 29.27 | 70.73 |  |
| Private rental | 4.3 | 45.23 | 54.77 |  | 31.26 | 68.74 |  |
| Other | 0.6 | 5.08 | 94.92 |  | 4.57 | 95.43 |  |
| Mean age (SD) | 48.6 (18.2) | 39.0 (13.7) | 51.6 (18.4) |  | 40.1 (15.7) | 49.6 (18.2) |  |
| General health, % |  |  |  | <0.001 |  |  | <0.001 |
| Excellent | 17.4 | 24.93 | 75.07 |  | 7.97 | 92.03 |  |
| Very good | 31.8 | 25.65 | 74.35 |  | 8.87 | 91.13 |  |
| Good | 28.1 | 24.65 | 75.35 |  | 10.94 | 89.06 |  |
| Fair | 15.1 | 21.77 | 78.23 |  | 12.72 | 87.28 |  |
| Poor | 7.6 | 20.31 | 79.69 |  | 13.85 | 86.15 |  |
| SF-12 Physical health, %<br>quartiles |  |  |  | <0.001 |  |  | <0.001 |
| 0-25% | 26.8 | 19.07 | 80.93 |  | 11.41 | 88.59 |  |
| 25-50% | 25.0 | 23.39 | 76.61 |  | 9.89 | 90.11 |  |
| 50-75% | 24.4 | 26.01 | 73.99 |  | 9.34 | 90.66 |  |
| 75-100% | 23.9 | 29.18 | 70.82 |  | 10.28 | 89.72 |  |
| SF-12 Mental health, %<br>quartiles |  |  |  | <0.001 |  |  | <0.001 |
| 0-25% | 24.4 | 28.59 | 71.41 |  | 15.35 | 84.65 |  |
| 25-50% | 24.5 | 25.99 | 74.01 |  | 9.65 | 90.35 |  |
| 50-75% | 26.2 | 23.08 | 76.92 |  | 7.91 | 92.09 |  |
| 75-100% | 24.8 | 19.51 | 80.49 |  | 8.32 | 91.68 |  |

### Housing and health outcomes: effect modification

Table 2 shows p-values for interaction terms of covariates sex, ethnicity, urbanicity and tenure, fitted separately for each covariate, health and unaffordability measure. For example, for all three health outcomes, Black and Asian participants showed different associations between housing affordability and health than white participants. As well, residing in housing that was mortgaged, socially rented or privately rented was associated with weaker housing unaffordability associations for all three health outcomes, except when using the 30% unaffordability measure with the mental health outcome (p-values for those with a mortgage, social renters, and private renters: 0.057, 0.019 and 0.052, respectively). Urbanicity only showed that rural participants had stronger associations of unaffordability with general and physical health, but not mental health, than urban participants when using the 30/40 unaffordability measure (p-values 30% vs 30/40: 0.114 vs 0.002 and 0.612 vs 0.044 respectively). Sex did not modify any associations between housing unaffordability and any health outcome.

**Table 2:** P-values from housing affordability^*^covariate interaction terms, fitted separately by covariate (n=35,114).

|  | Sex |  | Housing Tenure |  | Urbanicity |  | Ethnicity |  |  |
| --- | --- | --- | --- | --- | --- | --- | --- | --- | --- |
|  | - | Mortgage | Social Rental | Private Rental | - | White | Black | Asian | Other |
| General health, % |  |  |  |  |  |  |  |  |  |
| 30% measure | 0.507 | <0.001 | 0.015 | 0.601 | 0.114 | <0.001 | <0.001 | <0.001 | - |
| 30/40 measure | 0.608 | <0.001 | <0.001 | 0.088 | 0.002 | <0.001 | <0.001 | <0.001 | - |
| SF-12 Physical health, % quartiles |  |  |  |  |  |  |  |  |  |
| 30% measure | 0.781 | <0.001 | 0.386 | 0.946 | 0.612 | <0.001 | <0.001 | <0.001 | - |
| 30/40 measure | 0.310 | 0.004 | 0.003 | 0.702 | 0.044 | <0.001 | <0.001 | <0.001 | - |
| SF-12 Mental health, % quartiles |  |  |  |  |  |  |  |  |  |
| 30% measure | 0.654 | 0.035 | 0.045 | 0.943 | 0.140 | 0.057 | 0.019 | 0.052 | - |
| 30/40 measure | 0.444 | 0.007 | 0.001 | 0.759 | 0.535 | 0.013 | 0.001 | 0.005 | - |

Note: Reference group for Housing Tenure is participants with housing owned outright. Reference group for Ethnicity is White participants.

### Housing affordability and health outcomes

Table 3 shows associations between housing unaffordability and quartile decreases in health score, before and after addition of each covariate. For all three health measures, higher odds of housing unaffordability was associated with worse health (a quartile reduction in each health score), with larger strengths of association for the 30/40 than 30% measure. For example, the odds ratios (95% CI) for not good vs good general health were 1.22 (1.16-1.29) for the 30% measure and 1.88 (1.76-2.01) for the 30/40 measure. A similar pattern was seen for physical and mental health, albeit slightly lower strengths of association [30% vs 30/40 measure: 1.12 (1.06, 1.17) vs 1.63 (1.52, 1.74) and 1.26 (1.20, 1.32) vs 1.55 (1.44, 1.67)]. For each health measure, adjustment for ethnicity and tenure each increased strengths of association with housing unaffordability. The reverse is true for urbanicity, associations were reduced but still apparent. In sensitivity analysis, when the “Other” tenure category was removed, the associations for general and physical health were maintained, whereas mental health did not have a significant association.

**Table 3:** Adjusted associations of housing affordability measures with health metrics, Understanding Society 2009-11 (n=35,114).

|  | Model 1: Age-adjusted only |  | Model 2: Age & sex |  | Model 3: Age, ethnicity, unaffordable*ethnicity |  | Model 4: Age, urban, unaffordable*urban |  | Model 5: Age, tenure, unaffordable*tenure |  |
| --- | --- | --- | --- | --- | --- | --- | --- | --- | --- | --- |
|  | OR | 95% CI | OR | 95% CI | OR | 95% CI | OR | 95% CI | OR | 95% CI |
| General health, % |  |  |  |  |  |  |  |  |  |  |
| 30% measure | 1.22 | 1.16, 1.29 | 1.22 | 1.16, 1.29 | 1.27 | 1.20, 1.35 | 1.18 | 1.12, 1.25 | 12.29 | 6.26, 24.14 |
| 30/40 measure | 1.88 | 1.76, 2.01 | 1.88 | 1.76, 2.01 | 2.14 | 1.97, 2.33 | 1.77 | 1.64, 1.90 | 14.02 | 6.95, 28.28 |
| SF-12 Physical health, % quartiles |  |  |  |  |  |  |  |  |  |  |
| 30% measure | 1.12 | 1.06, 1.17 | 1.12 | 1.06, 1.17 | 1.12 | 1.06, 1.19 | 1.10 | 1.04, 1.16 | 17.12 | 5.56, 52.72 |
| 30/40 measure | 1.63 | 1.52, 1.74 | 1.62 | 1.52, 1.74 | 1.69 | 1.55, 1.84 | 1.56 | 1.45, 1.68 | 23.79 | 6.10, 92.85 |
| SF-12 Mental health, % quartiles |  |  |  |  |  |  |  |  |  |  |
| 30% measure | 1.26 | 1.20, 1.32 | 1.26 | 1.20, 1.32 | 1.28 | 1.21, 1.36 | 1.22 | 1.16, 1.29 | 4.23 | 1.12, 16.02 |
| 30/40 measure | 1.55 | 1.44, 1.67 | 1.54 | 1.43, 1.66 | 1.65 | 1.51, 1.80 | 1.52 | 1.41, 1.65 | 6.57 | 1.95, 22.20 |

**Table 4:** Health outcomes by Housing Tenure and Ethnicity, Understanding Society 2009-11.

|  | Housing Tenure |  |  |  |  |  |  | Ethnicity |  |  |  |  |
| --- | --- | --- | --- | --- | --- | --- | --- | --- | --- | --- | --- | --- |
|  | TOTAL<br>(n=35,114) | Owned<br>outright<br>(n=13,330) | Mortgage<br>(n=15,526) | Social<br>Rental<br>(n=4,551) | Private<br>Rental<br>(n=1,510) | Other<br>(n=197) | p-values<br>for<br>difference | White<br>(n=29,236) | Black<br>(n=1,686) | Asian<br>(n=3,652) | Other<br>(n=540) | p-values<br>for<br>difference |
| General health,<br>% |  |  |  |  |  |  | <0.001 |  |  |  |  | <0.001 |
| Excellent | 17.88 | 16.02 | 21.25 | 9.58 | 12.91 | 21.83 |  | 17.21 | 20.28 | 17.25 | 19.63 |  |
| Very good | 33.63 | 30.34 | 37.02 | 19.47 | 27.02 | 40.10 |  | 32.15 | 29.24 | 30.12 | 32.04 |  |
| Good | 28.51 | 28.63 | 27.61 | 26.72 | 31.99 | 23.35 |  | 27.89 | 27.28 | 29.98 | 25.93 |  |
| Fair | 13.59 | 17.20 | 10.59 | 23.91 | 17.88 | 9.64 |  | 15.25 | 16.13 | 14.05 | 12.96 |  |
| Poor | 6.39 | 7.81 | 3.53 | 20.33 | 10.20 | 5.08 |  | 7.50 | 7.06 | 8.60 | 9.44 |  |
| SF-12 Physical<br>health, %<br>quartiles |  |  |  |  |  |  | <0.001 |  |  |  |  | 0.007 |
| 0-25% | 21.04 | 33.39 | 15.53 | 45.24 | 28.74 | 22.34 |  | 27.11 | 23.96 | 25.38 | 26.11 |  |
| 25-50% | 24.11 | 26.81 | 24.63 | 21.29 | 23.77 | 25.89 |  | 24.97 | 25.21 | 25.41 | 22.96 |  |
| 50-75% | 26.15 | 22.31 | 28.75 | 15.86 | 23.25 | 25.89 |  | 24.32 | 24.67 | 25.11 | 22.04 |  |
| 75-100<br>% | 28.70 | 17.49 | 31.10 | 17.60 | 24.24 | 25.89 |  | 23.60 | 26.16 | 24.10 | 28.89 |  |
| SF-12 Mental<br>health, %<br>quartiles |  |  |  |  |  |  | <0.001 |  |  |  |  | <0.001 |
| 0-25% | 23.04 | 19.46 | 21.95 | 43.46 | 37.48 | 17.26 |  | 23.15 | 30.49 | 31.24 | 29.26 |  |
| 25-50% | 23.94 | 21.92 | 28.24 | 19.86 | 22.65 | 25.89 |  | 24.51 | 24.20 | 24.34 | 25.93 |  |
| 50-75% | 26.62 | 26.77 | 29.42 | 15.43 | 19.80 | 34.52 |  | 27.00 | 20.52 | 22.81 | 24.81 |  |
| 75-100<br>% | 26.40 | 31.85 | 20.39 | 21.25 | 20.07 | 22.34 |  | 25.34 | 24.79 | 21.60 | 20.00 |  |

**Table 5:**
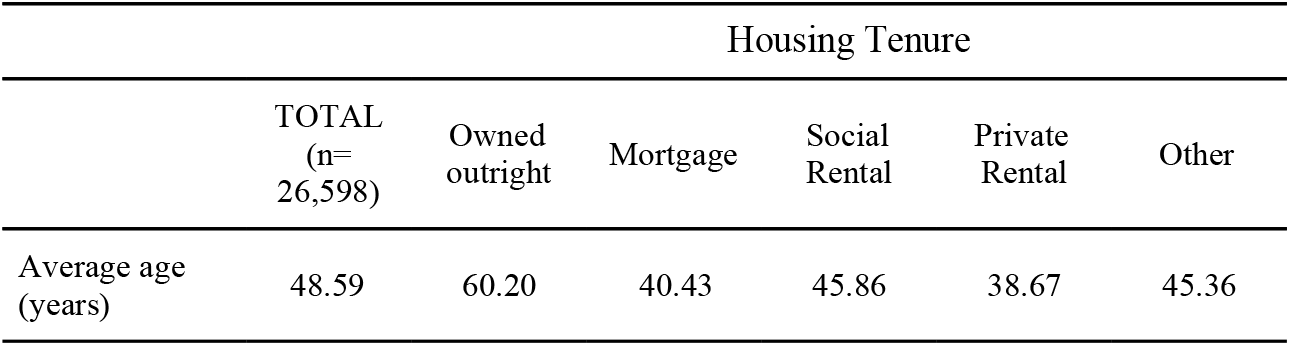
Average Age by Housing Tenure, Understanding Society 2009-11.

|  | Housing Tenure |  |  |  |  |  |
| --- | --- | --- | --- | --- | --- | --- |
|  | TOTAL<br>(n=<br>26,598) | Owned<br>outright | Mortgage | Social<br>Rental | Private<br>Rental | Other |
| Average age<br>(years) | 48.59 | 60.20 | 40.43 | 45.86 | 38.67 | 45.36 |

In models adjusted for age, sex and urbanicity, ethnicity and tenure were independent effect modifiers of relationships between housing unaffordability, in all three health outcomes. As tenure is the only modifiable factor of the two, figures 1-3 show odds ratios of a 1-quartile increase in a health measures for unaffordable compared to affordable housing, stratified by housing tenure and adjusted for age, sex, urbanicity, ethnicity and ethnicity^*^unaffordability. Physical, mental, and general health outcomes showed similar results, with mortgage owners and private renters reporting weaker associations between health and unaffordability. Associations were slightly stronger using the 30/40 compared to 30% income measure (Figures 1, 2, and 3). Social renters showed stronger associations with health than homeowners for general health (Figure 3). The physical health measure (Figure 1) was the same as the other two health measures, with the exception of no association between health and unaffordability in mortgage owners compared to homeowners using the 30% income measure.

**Figure 1.**
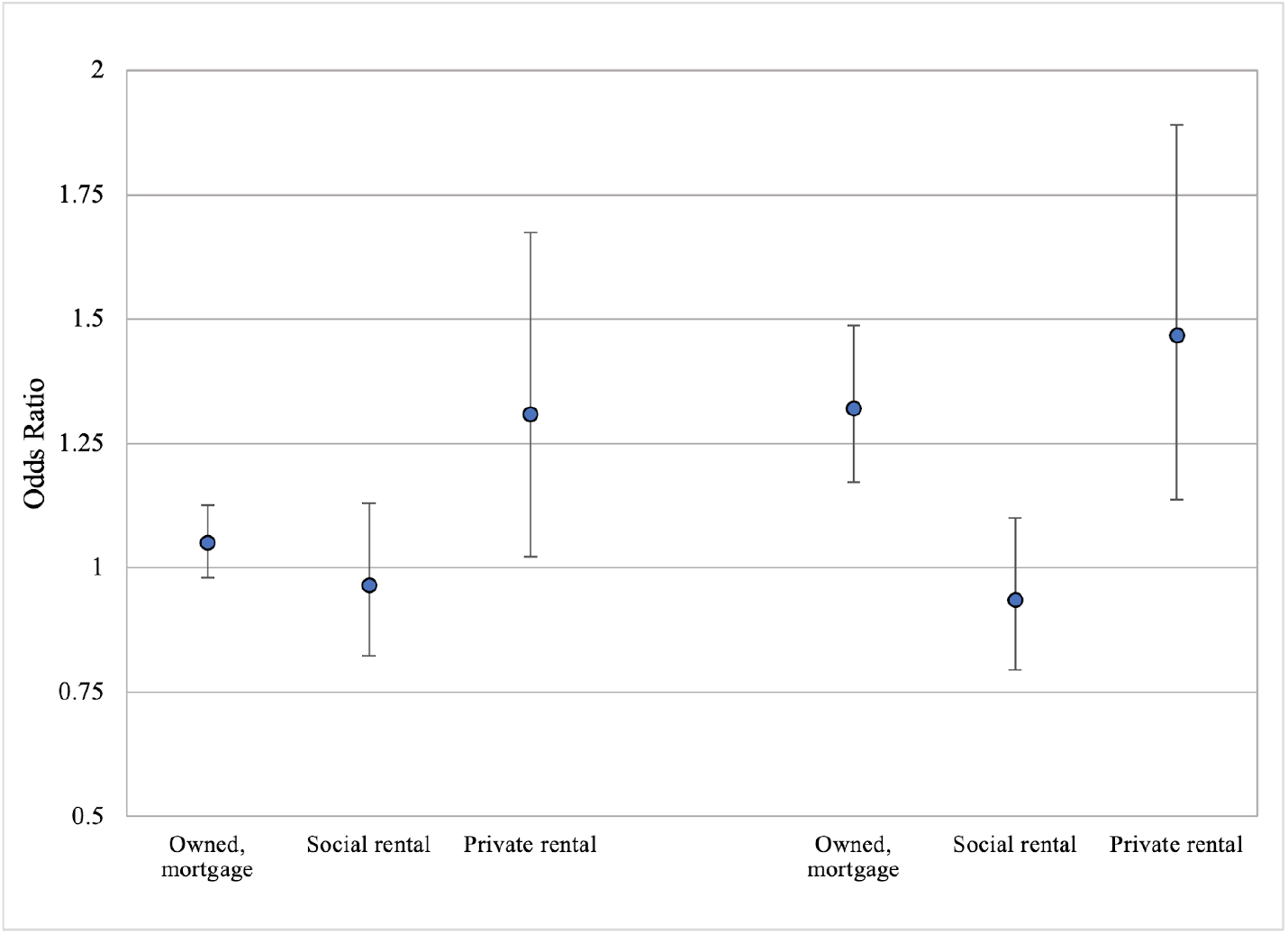
Adjusted^*^ odds ratios of a 1-quartile increase in physical health with unaffordable housing (left=30% measure, right=30/40 measure): stratified by tenure (n=35,114).

**Figure 2.**
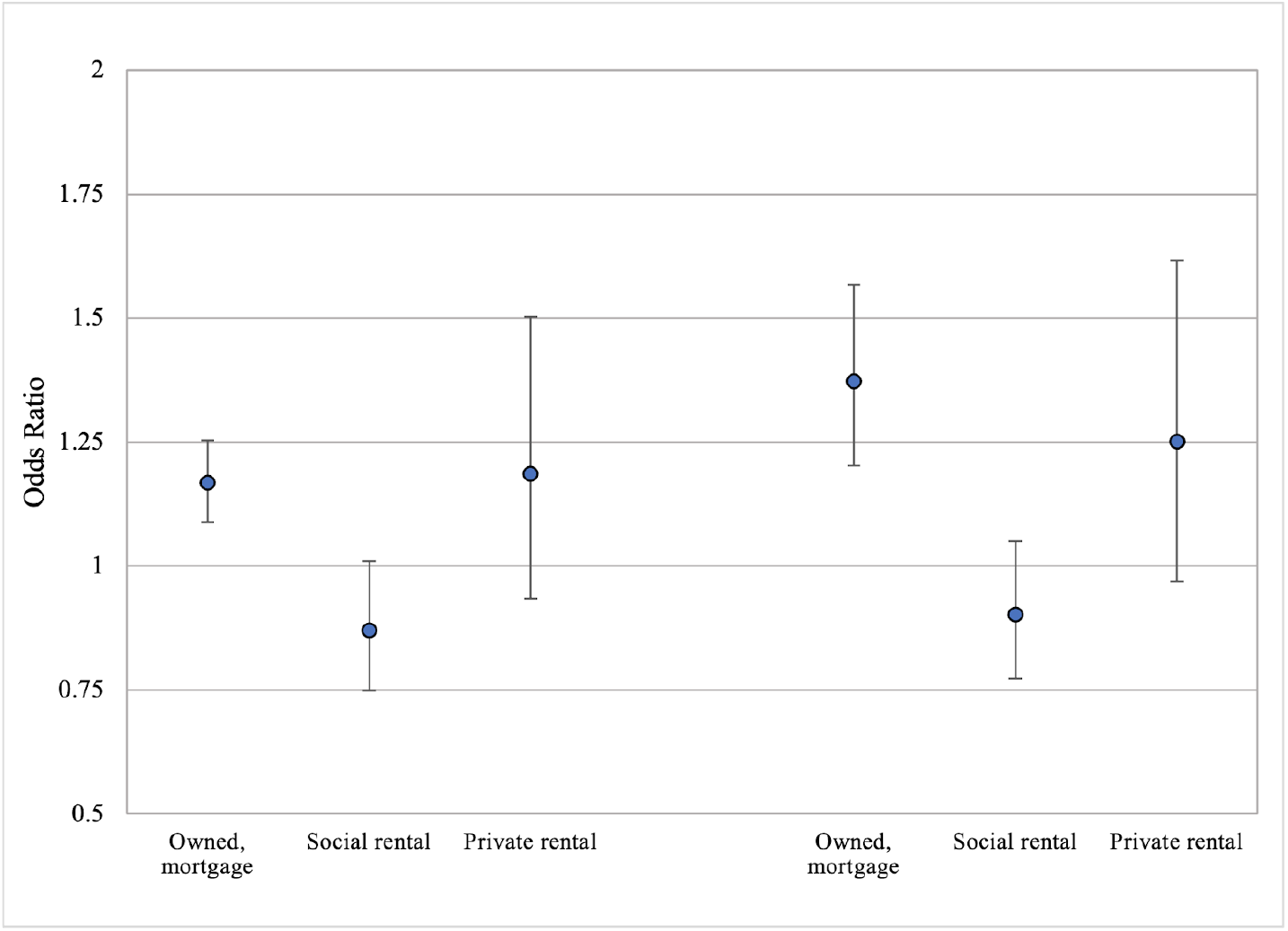
Adjusted^*^ odds ratios of a 1-quartile increase in mental health with unaffordable housing (left=30% measure, right=30/40 measure): stratified by tenure (n=35,114).

**Figure 3.**
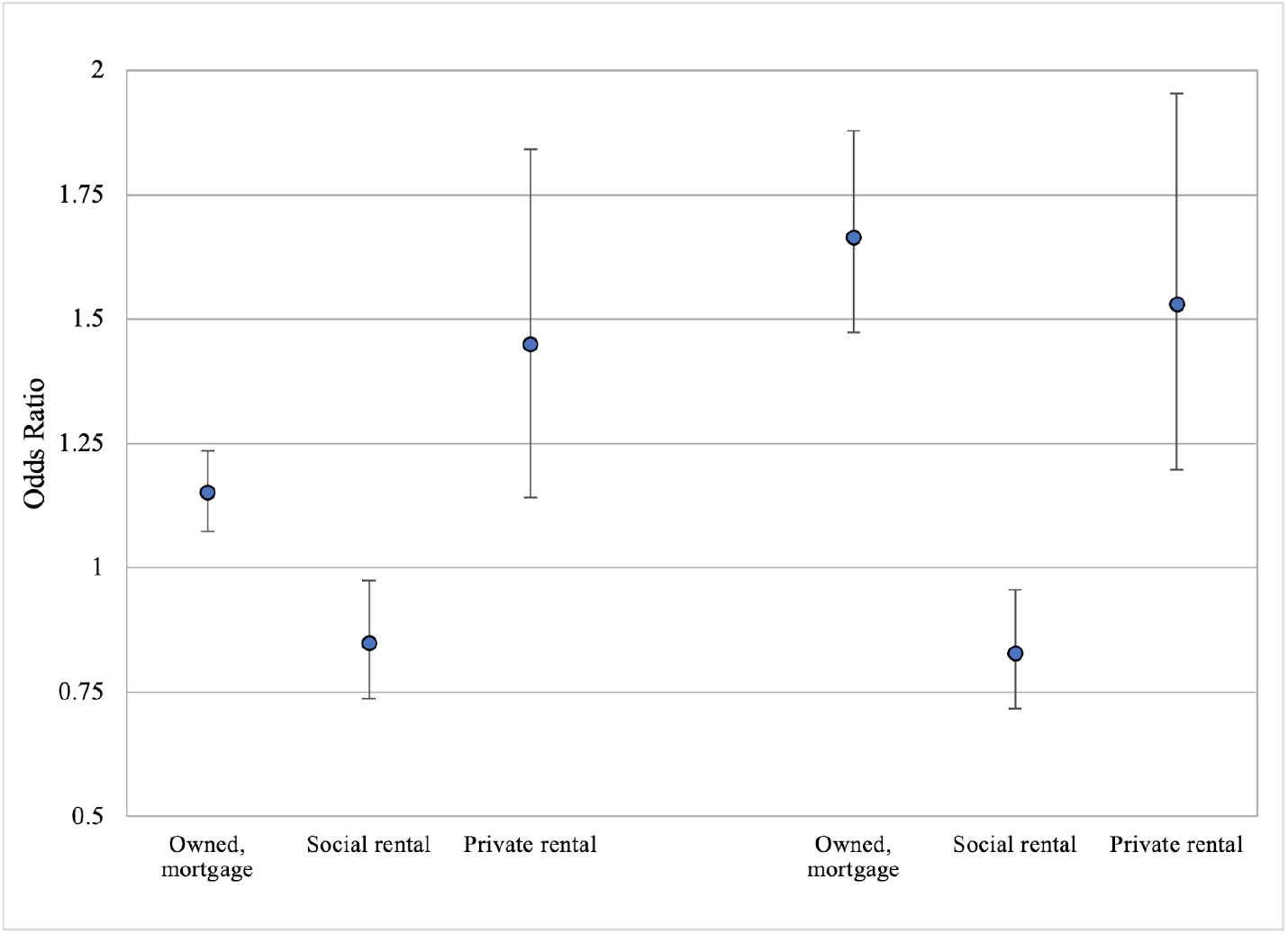
Adjusted^*^ odds ratios of a 1-quartile increase in general health with unaffordable housing (left=30% measure, right=30/40 measure): stratified by tenure (n=35,114).

## Discussion

### Summary of results

In this large population-based study on associations between housing affordability and different health metrics we showed a number of important findings. Firstly, whether using the 30% or 30/40 measure, living in unaffordable housing was associated with worse mental, physical and general health. However, importantly, the 30/40 measure demonstrated stronger relationships between unaffordability and worse health than the 30% measure. Second, and surprisingly, weaker associations were seen between housing affordability and health for some ethnic groups compared to White participants, and non-homeowner housing tenures compared to home-owners. Overall, results point to complex relationships between housing unaffordability, health, ethnicity and housing tenure.

### Comparison with other studies

Our overall finding that higher unaffordable housing is associated with worse health is consistent with other studies (Cairney & Boyle, 2004; Mason et al, 2013). However, previous studies examined one affordability measure only. We build upon these studies by showing that the 30/40 measure demonstrated a stronger association with health measures than the 30% affordability measure. This is likely due to the fact that the 30/40 metric is more effective at distinguishing between low and high income individuals, while 30% unaffordability also includes higher income individuals, who are typically in better health (Zaninotto et al., 2020). Past research has investigated the weaknesses of various housing affordability measures, including quantitative cross sectional and longitudinal analyses and qualitative literature reviews that looked at housing cost to income ratio (Rea, 2008; Stephen & Hoskara, 2019). Other studies have adjusted the housing cost to income ratio at different levels to better understand declining housing affordability in Western Europe (Dewilde, 2018). But as far as we are aware this is the first paper to compare multiple measures of housing unaffordability and how they vary for several outcomes.

The findings that minority participants experienced weaker associations between housing affordability and health for both affordability measures, has not been seen in the literature previously. This may be attributed to the small health disparities we observed between those with affordable and unaffordable housing among minority participants. In line with past studies that have identified ethnic minorities as vulnerable groups facing high rates of burdensome housing costs (Stone, 2004), in our data, a much larger proportion of Black, Asian and Other ethnic study members live in unaffordable housing than White study members, for both unaffordability measures. As well, patterns of health outcomes by ethnicity were less consistent in minority than White participants. For example, White participants had higher prevalence of worse physical health than minority study members. Another explanation may be that housing affordability has a greater share of influence on the health of White participants, while worse health outcomes among minority individuals may be a result of several issues faced, with housing affordability having less of an effect. More research is needed on this topic.

We also surprisingly found differences in directions of associations between housing unaffordability and health depending on tenure of the study member. In general, for all three health outcomes, private renters showed positive, while social renters showed no or inverse, associations with housing unaffordability. In our data, a similar proportion of private renters were categorized into unaffordable housing as mortgage owners (45.2% vs 40.7%), but the former were much more likely to be classified into the worst physical and mental health quartiles. Social renters were the opposite, with those in the affordable housing category demonstrating worse physical and mental health than home-owners. The reason mortgaged home owners only saw associations between housing affordability with the 30/40 measure is likely due to a higher proportion of people in the healthiest quartiles moving from being classified in ‘unaffordable’ housing with the 30% measure to ‘affordable’ housing with the 30/40 measure (data not shown).

### Study Strengths and weaknesses

This study had a number of strengths, Understanding Society (US) is one of the only studies with detailed financial data on household income, housing cost, demographics and health. This allowed us to analyze the sample for those suffering worse health when utilizing different definitions of housing affordability and health. Additionally, SF-36 scores have been validated and widely used to evaluate general and older population health (Gandek et al., 2004). The overall sample is also representative of the UK - for example the UK population has a similar proportion of Caucasian individuals to the sample (*Ethnicity and National Identity in England and Wales: 2011*, 2011). Coupled with the sample also having been oversampled for ethnic minorities, this allowed us to have large enough sub-samples to reliably test effect modification of associations by ethnicity. This is important, given that characteristics of sex, ethnicity, urbanicity, housing tenure, and age have demonstrated significant associations with housing affordability in past studies (Gan & Hill, 2009; OECD, 2011a; Cairney & Boyle, 2004; Rea W, 2008; Stone, 2004; Barton, 2017).

A number of study weaknesses should be acknowledged. A complete case analysis was done, with the sample analyzed having lower levels of 30% housing unaffordability, better mental health, and worse physical health compared to the excluded sample, and therefore may not be representative. Participants in the observed sample were older (average of 48.59 years old) than those excluded (average of 25.66 years old) and the general UK population (approximately 39 years old in 2011) (Office for National Statistics, 2018). Future studies could focus on distinct age groups, since physical health is intrinsically associated with age (Gandek et al., 2004).

The affordability measures focus on the financial aspect but provide minimal indication of health issues including whether people have sufficient diets, exercise enough, and are housed effectively from harmful conditions (Akbar & Hossain, 2017; Jones-Rounds et al., 2014; Rockwood and Mitnitski, 2020). As well spending 30% of income on housing as a threshold does not mean that some individuals spending less than 30% on housing are burden-less. Meanwhile, the 30/40 measure assumes all households above the 40th percentile of the income distribution are not facing housing unaffordability. For example, a household in the 39th percentile and a household in the 41st percentile spending thirty percent of income on housing may face similar financial issues. Only the family in the 39th percentile is considered as dealing with housing unaffordability. The metrics treat someone spending 31% of income on housing the same as spending 75% of income. It is a weakness that these financial measures do not consider the specific percent of income spent. This investigation sought to replicate measures that policymakers generally use, which is why a continuous measure of housing affordability was not applied.

Households may also under consume housing, where rent levels do not indicate overcrowding, or in the case of utilities, do not heat the living space to needed temperatures (Haffner & Boumeester, 2015). It has been recommended that nations utilize financial metrics centered around residual income, the money remaining after housing costs (Stone et al., 2011a). It focuses on whether people have enough money to consume all basic necessities and is applied by the U.S. Department of Veteran’s Affairs home loan program (Jewkes & Delgadillo, 2010). All other expenditures are adjusted to fit the remaining income, indicating that amount best determines when people are paying too much on housing (Stone, 2004). One issue with utilizing residual income to factor in is the complexity with analyzing it, since assumptions on income, taxes, and other benefits must be made (Stone et al., 2011b). Future work is needed to compare housing unaffordability and health associations with the residual income method.

General Health was self-reported by participants. It may be difficult to accurately compare health rates of different demographics, since there is a tendency to underreport information related to worse health (David W. Johnston, 2009).

Another limitation is the potential for reverse causation. Instead of housing unaffordability causing worse health, deteriorating health may worsen housing unaffordability. The cross-sectional analysis could be improved by doing a longitudinal analysis where health trajectories are measured. Lastly, covariates in this study may have demonstrated effect modification by chance. Power calculations were not run by sub-groups tested for effect modification, so there is a possibility that the power was lower in the stratified analysis.

### Recommendations for further research

Minority homeowners had significantly higher odds of poor health relative to White participants, yet had a weaker association between housing unaffordability and worse health. An investigation should be done to determine what factors contributed to this. Further analysis should be done to determine the extent that the introduction of affordable housing corresponds with improved physical and mental health outcomes and how each affordability metric responds. Lastly, investigations should be performed to determine if varying demographics have different health outcomes when presented with identical levels of housing unaffordability.

### Policy implications

#### Unitary and Dualist Housing Systems

Results of this analysis indicate an association between housing unaffordability and their impact on health. Policy approaches to improving health should reflect this understanding and focus on investments in increasing the housing supply as a way to improve health outcomes. Nations that have suffered from corporatized housing industries, like the UK and US, can shift from a dual system to a unitary model, where government supplied housing competes directly with the private sector (Norris & Byrne, 2018).

Unitary housing systems have been effective at creating affordable housing (Deutsch, 2009), such as in the city of Vienna, Austria (Marquardt & Glaser, 2020). The government of Vienna has spent decades purchasing land and constructing housing projects in order to bestow residents with affordable, public alternatives to the private market (Marquardt & Glaser, 2020). Over forty percent of households in Vienna live in social housing (Marquardt & Glaser, 2020). In contrast, less than one percent of US households live in public housing (HUD, 2021; U.S. Census Bureau, 2021). Seventeen percent of all housing in England is public housing (P. Chapman, 2019).

In nations with dual housing systems, access to social housing is limited to the poorest, leading to both resentment and negligible impact on overall housing affordability (Norris & Byrne, 2018; Van Duijne & Ronald, 2018). The cost advantage for providing demand-side subsidies to keep rents stable has waned in many cases, with a limited number of people receiving the support (Marquardt & Glaser, 2020; Apgar Jr., 2004). Restricted access and the limited subsidies on social housing protect the private industry from government competition (Norris & Byrne, 2018; Van Duijne & Ronald, 2018). Not only does private business benefit and profit from this setup, but the market gets less stable (Norris & Byrne, 2018). Homeownership is incentivized as renting becomes less and less affordable, resulting in more marginal homeowners with high risk of defaulting on mortgages. This drives extremely polarizing housing cycles (Mundt & Amann, 2009).

Unitary housing regimes work as a buffer against volatile prices, minimizing the damage from housing crises, such as in 2008 (Norris & Byrne, 2018). This can be seen when contrasting the situations in Ireland and Austria, countries with dual and unitary housing systems, respectively. In Ireland, social housing consists of 8.9% of total housing compared to 22% in Austria (Norris & Byrne, 2018). While Ireland restricted availability to low-income households, Austria allowed people of all income levels to utilize it (Marquardt & Glaser, 2020; Norris & Byrne, 2018; Van Duijne & Ronald, 2018). Furthermore, the private and public, non-profit rental sectors compete directly against each other, resulting in more stability (Amann, 2005; Norris & Byrne, 2018; Van Duijne & Ronald, 2018). Ireland has seen sinusoidal housing price changes, while Austria has experienced level prices (Norris & Byrne, 2018). The larger public market and commitment to building social housing resulted in Austria maintaining level prices, while Ireland’s reliance on a volatile dual system contributed to its housing crash in the late 2000s (Norris & Byrne, 2018).

In addition to stabilizing the market, unitary housing systems promote lower costs (Marquardt & Glaser, 2020; Mundt & Amann, 2009; Van Duijne & Ronald, 2018). In the UK, tenants spend greater than 30% of their income on rent (Rottier, 2018). Meanwhile, the housing expenditure rate in Austria is 18% (Amann, 2005). In Vienna specifically, a median income household spent approximately 13% of income on an average priced rental, not including running costs, in 2019 (Statistik Austria, 2021a; Statistik Austria, 2021b). This is a stark difference in expenditure levels between a nation that limits the influence of public housing offerings and a nation which has social housing compete in the market. When executed properly, social housing can be used as an apparatus against poverty (Amann, 2005).

As the social housing sector is diminished in size and influence, its ability to push down market rent levels dissipates, resulting in inflated costs and less affordable housing as seen in the UK. To reduce the population facing housing unaffordability by any metric, a focus on constructing social housing is advised.

## Conclusion

Of the two affordability measures analyzed, the 30/40 metric demonstrated a stronger association with health outcomes. It is recommended that nations like the U.S. and UK, which utilize the 30% affordability measure, re-evaluate how they define housing affordability and approach housing cost burden issues. Net income, factoring in housing costs, should be included in analyses that utilize cost ratios (ex. 30% and 30/40 metrics). Policy makers and researchers should consider expanding their definitions of unaffordability to more accurately identify populations vulnerable to adverse health effects of housing. Potential solutions focused on increasing the public housing supply can help people of various demographics confronted with housing unaffordability in nuanced ways. More research is needed to identify why housing unaffordability is more strongly associated with poor health in some housing tenures.

## Data Availability

The data set used in this study is available at www.understandingsociety.ac.uk.

https://www.understandingsociety.ac.uk/

## Data availability statement

The data set used in this study is available at www.understandingsociety.ac.uk.

